# The Network Landscape of Non-Clinical Eating Behaviors in India

**DOI:** 10.64898/2026.03.19.26348826

**Authors:** Dipanjan Ray, Anushka Ravishankar, Eunice Y Chen, Moumita Das

## Abstract

Eating behaviors are traditionally conceptualized as individual psychological phenomena, yet their structural organization remains largely unmapped in non-Western contexts. Adopting a dimensional perspective, this study used network analysis to characterize the architecture of non-clinical eating behaviors in India (N = 1,508; 48.9% male, 50.7% female; age range = 18-65 years) based on 35 well-validated biopsychosocial measures. Mixed Graphical Models revealed a highly optimized, small-world system defined by a dual-layered hierarchy. Cultural and living conditions (religion, home type) served as local behavioral anchors (highest expected influence), whereas socioeconomic factors (employment, education) and self-esteem functioned as structural bridges (highest betweenness centrality) integrating the biopsychosocial system. Notably, shape and weight concerns were highly predictable but operated as local cluster nodes rather than global integrators—directly challenging Western body-image-centric models. Limitations include cross-sectional design, precluding causal inference, and the need for ongoing validation of Western-derived measures in Indian contexts. These findings suggest that in a developing economy, broader psychosocial structural factors rather than individual factors may contribute to the maintenance of eating disorder symptoms.

## Introduction

Eating behaviors and their associated cognitive patterns have traditionally been examined within categorical, disorder-based diagnostic frameworks, wherein conditions such as anorexia nervosa or binge eating disorder are treated as discrete clinical entities. This “latent disease” model assumes that symptoms are passive indicators of an underlying common cause—a hidden pathogenic entity that “causes” the symptoms to co-occur Borsboom (2017). While this approach has yielded substantial insights into psychopathology, it often obscures the dimensional and continuous nature of eating-related experiences that extend well beyond diagnostic thresholds. This approach also struggles to explain high rates of comorbidity where symptoms overlap across diagnostic boundaries Robinaugh et al. (2020).

In response, an increasing body of scholarship has called for moving beyond categorical nosologies toward dimensional models that capture variation in symptoms and behaviors across both clinical and non-clinical populations Insel et al. (2010); Borsboom (2017). From this perspective, eating behaviors and concerns—such as restrained eating, emotional eating, shape concern, or overvaluation of weight—are understood as existing along continua, frequently manifesting in subclinical forms that nonetheless carry meaningful psychological and health-related consequences.

Crucially, the empirical foundations of these dimensional models are overwhelmingly derived from Western, Educated, Industrialized, Rich, and Democratic (WEIRD) populations ^1^. This “universalist” bias assumes that the drivers of eating pathology—primarily internalized thin-ideals and body dissatisfaction—function identically across the globe. However, this assumption remains largely untested in the Global South ^2^.

India’s relevance to this question extends beyond its underexplored status. As the world’s most populous nation and among its fastest-growing major economies, India juxtaposes co-occurring extremes—persistent undernutrition alongside rising obesity, and rapid technological and economic advancement alongside traditional rural livelihoods—within a single population. National surveillance estimates indicate that underweight and obesity now coexist at substantial rates within the same country and often the same communities, making India an unusually stringent test case for models of eating behavior developed in more socioeconomically homogenous WEIRD samples. Embedded within a diverse matrix of sociocultural practices, religious norms, caste-linked food rules, and region-specific dietary traditions, eating in India is not merely a physiological necessity but a deeply social and symbolic act Harper (1961). This complexity is especially relevant for network approaches to psychopathology, which conceptualize disorders as emerging from interactions among symptoms and their environmental drivers rather than as expressions of a single underlying condition.

The network perspective on mental health is an emerging framework that conceptualizes psychological phenomena as complex biopsychosocial systems rather than as expressions of latent disorders Borsboom and Cramer (2013); Fried et al. (2017); Ebrahimi (2023). Within this framework, mental health conditions are understood as networks of mutually interacting behaviors, cognitions, and traits, whose interdependencies give rise to patterns of vulnerability and resilience. This perspective reflects a broader shift in mental health research toward recognizing that psychological difficulties arise not from isolated symptoms or singular causal pathways, but from dynamic interactions among biological, psychological, and social components. Such an approach is particularly well suited to identify whether the “core” of eating behavior in a non-Western population is anchored by different systemic drivers.

In the present study, we employ a Mixed Graphical Model (MGM) Haslbeck and Waldorp (2020); Fried et al. (2020), a statistical network approach designed to accommodate variables of mixed measurement types (continuous, categorical, and count data), to examine individual-level and broader social factors interacting to shape eating behavior in a large non-clinical Indian sample. We conceptualize anthropometric characteristics (e.g., self-reported BMI and health conditions), psychological factors (e.g., self-esteem, loneliness), demographic factors (e.g., regional variation, urban–rural residence, caste), social influences (e.g., peer pressure, family norms), and eating-specific cognitions and behaviors (e.g., binge eating, weight concern, purging, overexercise) as nodes within an interconnected system. Estimating conditional dependencies among these nodes allows us to construct a weighted network that reveals central features, clustering patterns, and cross-domain pathways linking biological, psychological, and social processes.

The goals of the present study were threefold: (1) to map the interrelations among biopsychosocial variables and eating-related constructs in a diverse Indian population; (2) to identify communities of variables that cluster together within the network; and (3) to determine central or “hub” nodes that may serve as leverage points for early intervention. Ultimately, this study seeks to shift the global discourse from a focus on downstream symptoms to the upstream structural factors ^3^ that define the landscape of eating behavior in the Global South.

## Research Transparency Statement

### General Disclosures

#### Artificial Intelligence

Generative AI (ChatGPT, OpenAI, model GPT-4) was used to assist with language refinement, copyediting, and word count reduction of the abstract to meet journal formatting requirements. The authors reviewed, fact-checked, and approved all final content. No AI was used for data analysis, figure generation, or conceptual development of the research.

#### Ethics Approval

This research received ethical approval from the Institutional Review Board (IRB) of Ashoka University.

### Study Disclosures

#### Preregistration

No aspects of the study were preregistered.

#### Data, Code, and Materials

All survey instruments, de-identified datasets, and analysis scripts are available as detailed in the *Data Accessibility Statement*.

## Method

### Participants

Participants (N = 1,508) were recruited from a geographically diverse Indian cohort via an online panel (Zoho Survey). Of the initial 1,791 respondents, 283 (15.8%) were excluded for failing at least one of four attention checks embedded throughout the survey. The final analytical sample thus comprised 1,508 participants. The sample exhibited a balanced gender distribution (48.9% Male, 50.7% Female). Consistent with the digital nature of the recruitment platform, a substantial majority of respondents resided in urban areas (86.4%). Geographically, the sample covered a wide range of Indian states and Union Territories, with the highest representation from Maharashtra (12.9%), Kerala (11.5%), and Delhi (9.4%). Regarding socioeconomic status, the majority of participants (61.5%) identified as General Category, followed by Other Backward Class (22.2%) ^4^ These categories serve as critical proxies for social stratification in the Indian context. In terms of religious affiliation, the sample was predominantly Hindu (74.8%) and Muslim (13.0%). Educationally, the largest segment of the sample held an Undergraduate degree (45.0%), while 29.2% possessed a Postgraduate degree. A detailed summary of the socio-demographic and economic characteristics in Figure 1.

**Figure 1:**
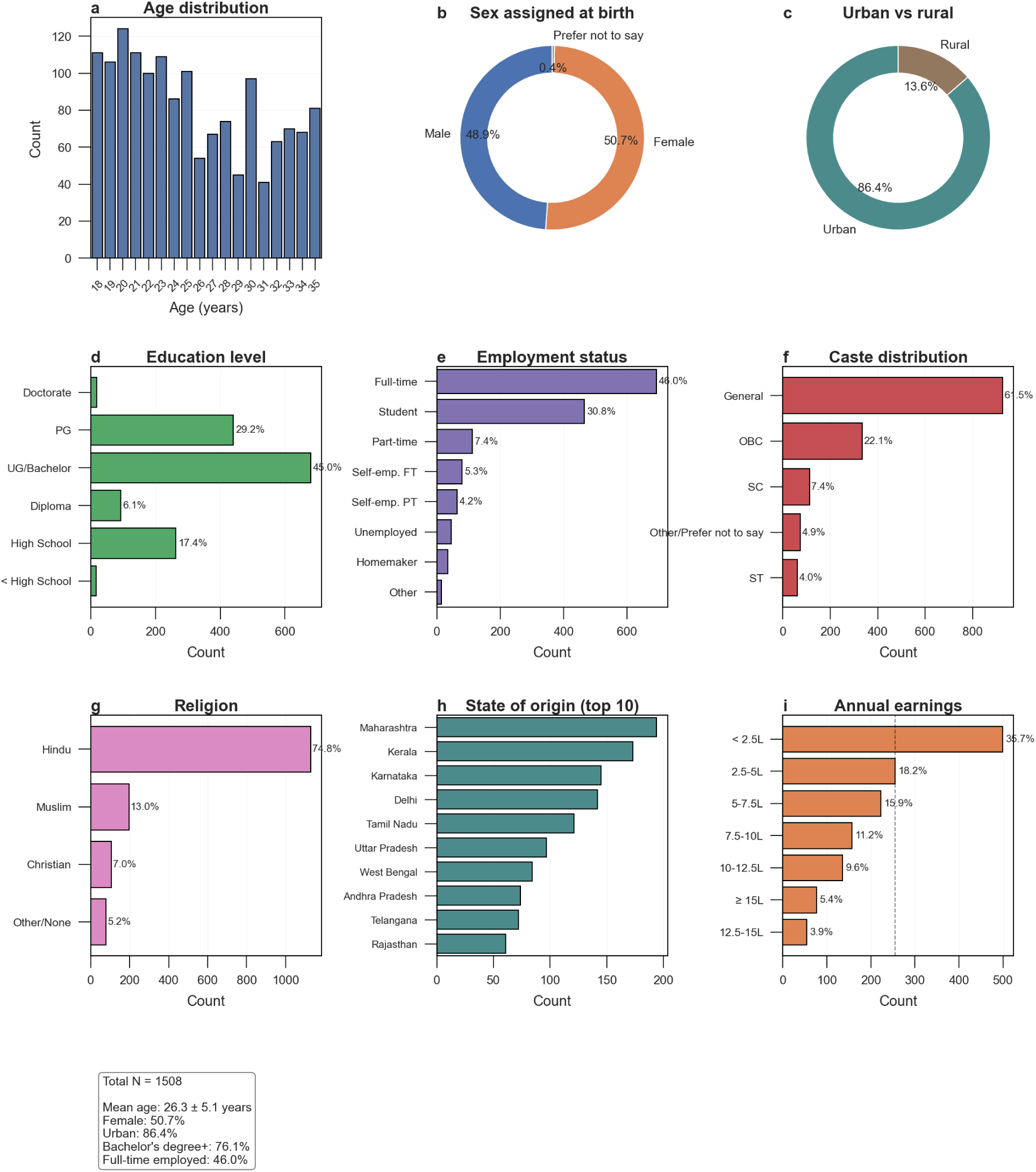
Demographic Overview of Participants

### Sample Size Justification

The study sample (*N* = 1, 508) was determined to provide sufficient power and stability for the estimation of a 35-node Mixed Graphical Model (MGM). For an undirected network of this complexity, there are *p*(*p −* 1)/2 = 595 potential pairwise interaction parameters. While early guidelines suggested a minimum ratio of three observations per parameter (*N* ≈ 1, 785; (Fried and Cramer, 2017)), our analytical framework ensures structural integrity at the current sample size through three primary mechanisms:

- **Regularization via 10-fold Cross-Validation:** To manage the parameter-to-observation ratio (approx. 1:2.5) and minimize Type 1 errors, we employed the Least Absolute Shrinkage and Selection Operator (LASSO). Unlike fixed heuristics, we utilized 10-fold cross-validation to empirically select the tuning parameter (*λ* ) that minimized the objective function. This approach ensures that the model only retains edges with genuine predictive utility, effectively shrinking the parameter space and mitigating the risk of overfitting (Haslbeck and Waldorp, 2020).
- **Empirical Stability Metrics:** The adequacy of the sample size was verified post-hoc using non-parametric bootstrapping (1,000 iterations). We calculated the Centrality Stability (CS) coefficient to ensure that the relative rankings of node importance (Expected Influence and Betweenness) were robust to sampling variation. Following established benchmarks (Epskamp et al., 2018), a CS-coefficient exceeding 0.50 was considered indicative of high stability, confirming that the network architecture is reliably estimated.
- **Contextual Power:** A sample of *N* = 1, 508 represents one of the largest biopsychosocial networks mapped in a non-Western, non-clinical context. This sample provides significantly higher sensitivity for detecting low-weight edges compared to the median sample sizes reported in contemporary psychopathological network literature (typically *N* < 500), allowing for the identification of the critical ”bridge nodes” central to our findings.

### Ethics and Data Integrity

The study protocol received ethical approval from the Institutional Review Board of the authors’ university (IRB Protocol ID anonymized for review). All participants provided informed electronic consent prior to enrollment, with the option to withdraw at any point without penalty. The cohort was screened for non-clinical status, excluding individuals with self-reported history of psychiatric or neurological disorders.

To ensure high data fidelity, we implemented a rigorous four-stage attention-check protocol designed to identify careless, inattentive, or automated responding. Participants who failed any single attention check (n = 283; 15.8% of initial respondents) were immediately excluded from the final sample. The four attention checks consisted of:

1. **Instructed response item 1:** ”Have you been paying attention to each question? If yes, select six i.e., everyday.”
2. **Factual knowledge check:** ”How many sides does a square have?” (Correct answer: 4 sides.)
3. **Instructed response item 2:** ”If you are paying attention, select five i.e., strongly agree.”
4. **Instructed response item 3:** ”When you have read this sentence, select option two i.e., sometimes.”

This multi-stage protocol was designed to identify participants who were not carefully reading instructions or responding systematically, as such careless responding can introduce noise and bias into network estimation.

Following application of these exclusion criteria, the final analytical sample comprised N = 1,508 participants.

### Online Data Collection and Response Quality Assurance

We implemented comprehensive procedures to ensure response quality and detect careless, inattentive, or automated participation.

#### Prevention of duplicate and automated submissions

Upon survey initiation, participants completed a CAPTCHA verification to deter automated scripts. IP address filtering was applied to identify potential duplicate submissions. While some IP addresses repeated (maximum five responses from a single IP), this pattern is consistent with natural sharing of WiFi networks, college campus environments, and office settings with no evidence of systematic flooding from a single source.

#### Attention and carelessness screening

As detailed above, four attention checks were embedded throughout the survey at approximately evenly spaced intervals. These included both instructed response items (e.g., “select option two”) and factual verification questions (e.g., “How many sides does a square have?”). This design allowed us to identify participants who were not reading instructions carefully or who were responding randomly. The use of multiple attention checks distributed across the survey length reduced the likelihood that inattentive participants would pass all checks by chance.

#### Response time screening

Completion times were monitored to identify implausibly fast responses indicative of careless clicking. Completion times exhibited a median of approximately 20 minutes and a mean of 24 minutes, suggesting that most participants engaged thoughtfully with the survey. The fastest completion time was approximately 5.1 minutes; while rapid, this remained within plausible bounds for conscientious responding by a very fast reader. A long tail extending to several hours was observed, likely reflecting respondents who paused the survey or left it open while completing other tasks. No participants were excluded solely based on response time, as timing alone can be an unreliable indicator of carelessness; however, extremely fast completions (under 5 minutes) in combination with attention check failures were captured by the attention check protocol.

#### Open-text response scrutiny

Free-text responses were manually reviewed for evidence of careless or automated generation. Text entries demonstrated natural linguistic variability, contextually appropriate content, and human-typical idiosyncrasies (e.g., colloquialisms, varied sentence structures, occasional typographical errors, and personalized responses). Importantly, no open-text responses contained gibberish, repetitive characters, or clearly nonsensical content that would indicate random clicking or automated generation.

#### Data quality inspection and preprocessing

Post-hoc data inspection revealed patterns characteristic of human—rather than automated or perfectly consistent—responding. Specifically:

- **Height field heterogeneity:** The survey requested height in feet and inches. However, many respondents entered values in centimeters, decimal feet, or mixed units—a pattern typical of human misunderstanding or conversion attempts rather than systematic automated responses.
- **Plausible outlier detection:** One extreme outlier in the compulsive exercise measure (Q18) reported 160,000 episodes in the past 28 days—a biologically and logically impossible value (equivalent to approximately 5,714 episodes per day). This value was determined to be a data entry error (likely a typo or unit misunderstanding) and was winsorized during preprocessing. The presence of such improbable but human-typical entry errors, rather than perfectly clean or systematically patterned data, further supports the authenticity of human participation.

#### Summary

The combination of CAPTCHA verification, IP analysis, multiple distributed attention checks, response time monitoring, manual open-text review, and post-hoc data quality inspection supports the integrity of the final sample (N = 1,508). We find no evidence of systematic automated responses, and the rate of careless responding (15.8% exclusion) is within expected ranges for online survey research with attentive participants.

### Measures

Thirty-five variables spanning demographic, biological, psychological, social, and eating-related domains were included. (see Table 1 for complete list and rationale).Detailed information about all measures, including the EDE-Q subscales, DERS-16, FMPS, RSES-10, DJGS, SATAQ-4, and HFIAS, along with their psychometric properties, is provided in Supplementary Material Section I.a (Measures).

**Table 1:** Rationale for inclusion of nodes in the network analysis.

| Node | Rationale | Key References |
| --- | --- | --- |
| Age, Gender, Region, Urban/Rural, Caste, Religion | Basic demographic variables linked to variability in eating patterns and social determinants of health | (Vaidyanathan and Menon, 2024; Jain et al., 2024) |
| Education, Employment, Family Income, Home Type | Socioeconomic indicators often associated with food access, dietary quality, and psychological distress | (Darmon and Drewnowski, 2008; Bhargava, 2004) |
| BMI, Health Condition | Indicators of physical health status and metabolic risk, closely related to eating patterns and body image | (Bulik and Reichborn-Kjennerud, 2003; Arcan et al., 2014) |
| Parent History of Eating Disorder | Genetic/familial predisposition and intergenerational transmission of disordered eating | (Strober et al., 2000; Lilenfeld et al., 1998) |
| Concern/Doubt, Personal Standards, Loneliness, Emotion Regulation, Self-Esteem | Psychological variables shown to influence disordered eating via perfectionism, emotional dysregulation, and self-concept | (Mora et al., 2017; Arcelus et al., 2013, 2015; Fairburn et al., 2003b; Levine, 2012; Wilson, 2017) |
| Food Insecurity | Chronic stressor associated with disordered eating and binge behaviors in multiple populations | (Becker et al., 2017; Hazzard et al., 2020) |
| Family, Peer, Media Pressure | Key sociocultural factors contributing to body dissatisfaction and dietary control attempts | (Rodgers et al., 2021; Levine and Murnen, 2009) |
| Diet Type, Food Source, Diet Change in Past | Dietary behavior indicators reflecting nutritional intake patterns and behavioral flexibility | (Mörkl et al., 2024; Nasser, 2009; Bardone-Cone et al., 2012; Story et al., 2008) |
| Overeating, Loss of Control, Binge Days, Purging, Overexercise | Core behavioral markers of disordered eating patterns; widely used in both clinical and dimensional frameworks | (Hudson et al., 2007; Keel, 2017) |

**Note.** *Personal Standards* and *Concern/Doubt* capture perfectionism dimensions regarding setting unrealistically high personal performance goals versus worrying over errors and self-doubt; *Emotion Regulation* reflects difficulty in understanding, accepting, and managing negative emotional states; *Self-Esteem* measures global feelings of self-worth and self-acceptance; *Loneliness* assesses perceived social isolation and emotional connectedness; *Family, Peer, and Media Pressure* capture perceived external pressure to conform to thin or appearance ideals; *Food Insecurity* indicates limited or uncertain economic and physical access to sufficient food; *Health Condition* refers to self-rated overall physical health status (ranging from Excellent to Poor); *Home Type* indicates self-reported dwelling structure; *Food Source* denotes participants' primary reported mode of household food procurement or daily preparation (e.g., market purchase, home-grown, canteen, subsidized ration); and behavioral nodes (*Overeating, Loss of Control, Binge Days, Purging, Overexercise*) capture specific disordered eating frequency markers over the past 28 days (e.g., consuming large food quantities, feeling unable to stop eating, and compensatory practices). Comprehensive details on all measurement tools, item structures, and operational definitions are available in the Supplementary Document.

### Analytical Framework: Mixed Graphical Models (MGM)

To investigate the conditional dependencies between variables of mixed measurement types (Gaussian, categorical, and count), we implemented a Mixed Graphical Model (MGM) using the mgm package in R (version 4.2.0). The MGM estimates the joint probability distribution by combining different exponential families. Pairwise interaction potentials represent the conditional dependency between two nodes, controlling for all other variables in the system (Haslbeck and Waldorp, 2020). The details of the network measures used in the analysis can be found in Table 2.

**Table 2:** Network Measures Used in the Analysis. Standard formulations and interpretations for these metrics are derived from established methodologies in network psychometrics and graph theory (Haslbeck and Waldorp, 2018; Watts and Strogatz, 1998; Humphries and Gurney, 2008; Robinaugh et al., 2016; Freeman, 1977; Blondel et al., 2008).

| Measure | Description and Relevance |
| --- | --- |
| *Network Model Estimation* |  |
| **Edge Weight (MGM)** | Represents the conditional association (partial correlation) between two variables, controlling for all other variables in the network. Estimated for mixed data types (Gaussian, categorical). |
| **Node Predictability ($R^2$)** | The proportion of a node’s variance that is explained by its direct neighbors. Indicates how strongly a node is embedded and influenced by the network. |
| *Global Topology* |  |
| **Global Clustering Coefficient** | The overall tendency of nodes to form clusters. A high value indicates a network with tightly connected local neighborhoods. |
| **Average Path Length** | The average number of steps along the shortest paths for all possible pairs of network nodes. A low value indicates high global efficiency. |
| **Small-worldness Index** | A ratio of the network’s clustering coefficient and path length relative to random networks. Values $> 1$ indicate a "small-world" structure. |
| *Node-level (Integration)* |  |
| **Expected Influence (EI)** | The sum of all edge weights (both positive and negative) connected to a node. Identifies nodes expected to have the strongest activating or inhibiting effect on the system. |
| **Betweenness Centrality** | Number of shortest paths that pass through a node. Identifies critical "bridge" or "broker" nodes that connect disparate communities. |
| *Meso-level (Segregation)* |  |
| **Community Detection (Louvain)** | A modularity-maximization algorithm that iteratively groups nodes to find a community structure with the highest modularity score. |

#### Regularization

To ensure model sparsity and minimize Type I errors, we employed the Least Absolute Shrinkage and Selection Operator (LASSO). The optimal tuning parameter (*λ* ) was selected via 10-fold cross-validation. We applied the ’OR’ rule for edge inclusion, where an edge is retained if a dependency is detected in either neighborhood regression. The formal mathematical specification of the Mixed Graphical Model (MGM), including the nodewise estimation procedure with *ℓ*_1_-regularization and *K*-fold cross-validation for selecting the tuning parameter, is described in Supplementary Material Section I.b (Formal Mathematical Specification of the MGM).

#### Node predictability

The predictability of each node indicates the proportion of its variance that is explained by its direct neighbors in the network. We quantified node predictability using *R*^2^ for continuous variables and Classification Correctness (CC) for categorical variables (Haslbeck and Waldorp, 2018). Node predictability is detailed in Supplementary Material Section I.c (Node Predictability Measures).

### Network Stability

To assess the reliability of the estimated network structure and edge weights, a non-parametric bootstrapping procedure (1,000 iterations) was conducted. This analysis identifies the “robust backbone” of the biopsychosocial system by filtering out potentially spurious or sample-dependent associations. We calculated 95% confidence intervals for edge weights and the Centrality Stability (CS) coefficient using the bootnet package (Epskamp et al., 2018). Supplementary Material Section I.f (Data Pre-processing and Stability) describes the pre-processing steps, including Winsorization of behavioral variables and outlier handling, along with the non-parametric bootstrap procedure used to compute Centrality Stability coefficients.

### Global Topology and Small-Worldness

The global topological properties of the weighted network were quantified to characterize the overall system architecture. We characterized the global architecture using the Small-Worldness Index (S), which identifies systems that combine high local clustering with short global communication distances (Humphries and Gurney, 2008):

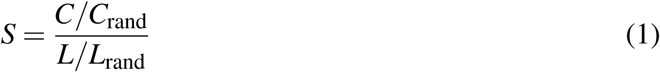

where C is the global clustering coefficient and L is the average weighted distance. Benchmark values (*C*_rand_, *L*_rand_) were generated from 100 degree-preserved random graphs.

These measures provide a mathematical snapshot of how efficiently information—or “pathological cascades”—can spread through the biopsychosocial system. The key metrics are summarized in Table 3.

**Table 3:** Global Network Topology Measures.

| Measure | Value |
| --- | --- |
| Average Weighted Distance ($L$) | 0.0297 |
| Weighted Diameter | 0.0877 |
| Assortativity (by Degree) | -0.1428 |
| Global Clustering Coefficient ($C$) | 0.5830 |
| Small-worldness Index ($S$) | 54.636 |

*Note:* $L$ and Diameter represent the average and maximum weighted shortest paths between nodes. Small-worldness ( $S$ ) was calculated as the ratio of the normalized clustering coefficient to the normalized path length relative to 100 degree-preserved random networks.

### Centrality Measures

To identify the most influential and “integrative” nodes within the network, we computed standardized centrality measures like Expected Influence (EI) and Betweenness centrality. EI represents the cumulative structural importance of a node by accounting for the sum of its shared variance with all neighbors. Betweenness centrality quantifies the degree to which a node lies on the shortest path between other node pairs, identifying the critical “bridges” that integrate disparate modules. These metrics distinguish between nodes that exert the most local influence and those that act as global “brokers” across the biopsychosocial system (Robinaugh et al., 2016; Freeman, 1977). Supplementary Material Section I.d (Network Topology and Centrality) provides the complete mathematical formulations for Expected Influence and Betweenness centrality, including the justification for using EI over traditional strength centrality in networks containing negative edges.

### Community Detection

To investigate the structural “segregation” of the network and identify densely interconnected modules, we employed the Louvain community detection algorithm (Blondel et al., 2008). The Louvain community detection algorithm, including the modularity maximization procedure and the two-phase iterative process, is formally specified in Supplementary Material Section I.e (Community Detection: Louvain Algorithm).

## Results

### Weighted Mixed Graphical Model (MGM) Analysis

The weighted MGM revealed a dense network of conditional dependencies among the 35 variables (Figure 2).

**Figure 2:**
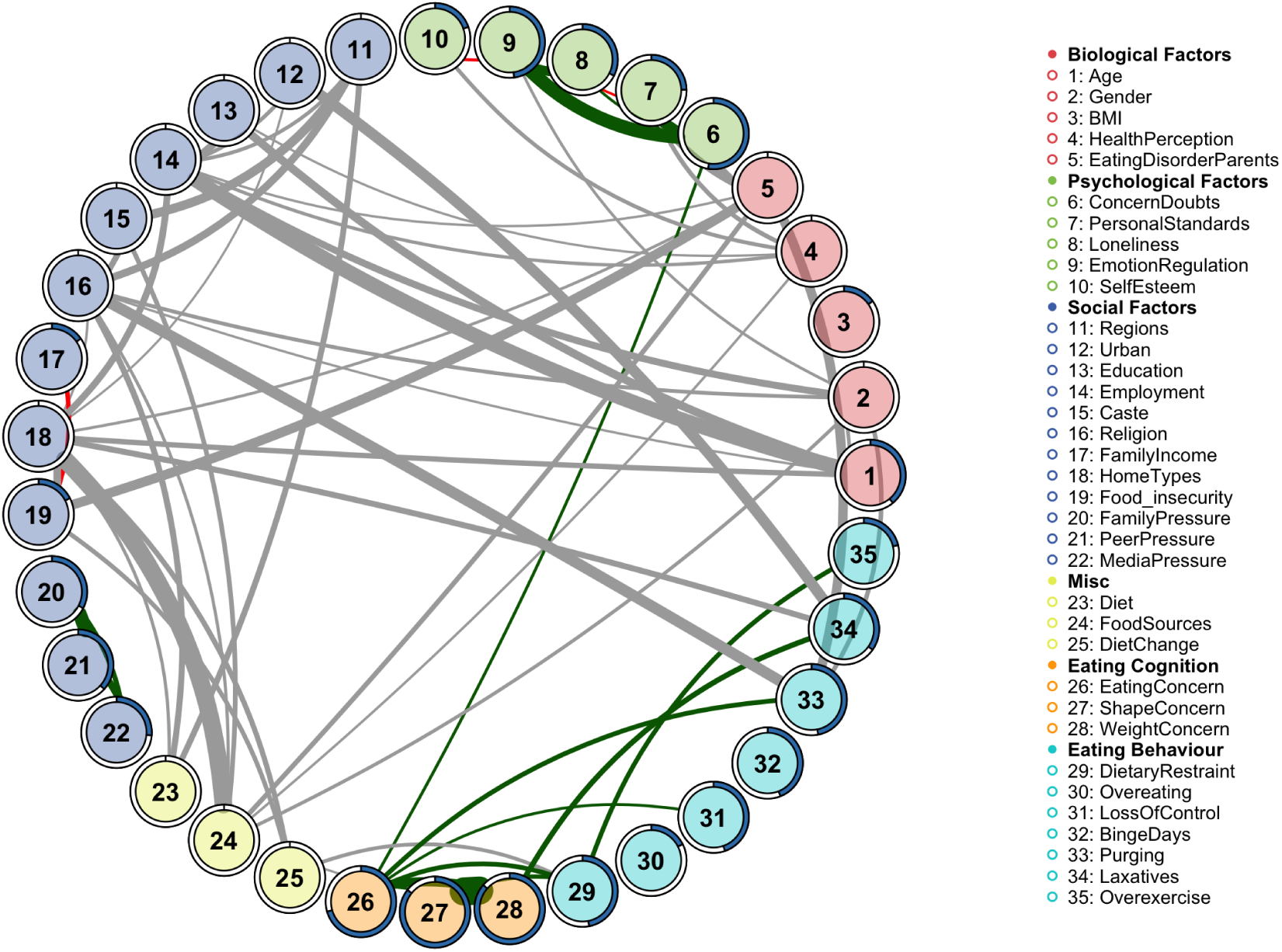
The Weighted Biopsychosocial Network of Non-Clinical Eating Cognition and Behavior: The network consists of 35 nodes, representing variables from biological (red), psychological (green), social (blue), miscellaneous (light blue), eating cognition (yellow), and eating behavior (orange) domains. Edges represent conditional dependencies. Green edges indicate positive associations, red edges (not prominently visible at this level) indicate negative associations, and grey edges represent non-valence associations (e.g., between categorical variables). Edge thickness is proportional to the association’s strength (weight). The pie fill within each node represents its predictability (*R*^2^), indicating the proportion of variance explained by its neighboring nodes. Variable names used in Results, tables, and figures follow an abbreviated coding convention for clarity in network visualizations.

#### Node Predictability

The predictability (*R*^2^) of each node, visualized in Figure 2 by the pie-fill (i.e., by the proportion of each node’s circular icon that is shaded, corresponding to its predictability, R², as defined in the Figure 2 caption) showed a clear pattern: nodes within the **Eating Cognition** (e.g., `EatingConcern` (26),`ShapeConcern` (27), `WeightConcern` (28)) and **Eating Behaviour** (e.g., `DietaryRestraint` (29),`LossOfControl` (31),`BingeDays` (32),`Purging` (33)) domains exhibited the highest predictability values. Psychological factors, such as `ConcernDoubts` (6), and `EmotionRegulation` (9) (assessed via the Difficulties in Emotion Regulation Scale, DERS-16; higher scores reflect greater emotion dysregulation) also showed high predictability. In contrast, many demographic and social factors (e.g., Gender (2), HealthPerception (4), Education (13), Employment (14), Caste (15), Religion (16)) had lower *R*^2^ values.

#### Network Edge Associations

The network edges, representing the strength and nature of conditional dependencies between variables, were estimated via the weighted MGM (Figure 2). The associations are categorized by their valence and measurement type:

- **Positive Associations (Green Edges):** Prominent positive dependencies were observed among several groups of nodes. Within the psychological domain, strong edges linked `ConcernDoubts` (6), `PersonalStandards` (7), `Loneliness` (8), and `EmotionRegulation` (9). In the eating-related domain, dense connectivity was found between cognitive variables—`EatingConcern` (26), `ShapeConcern` (27), and `WeightConcern` (28)—and behavioral variables including `DietaryRestraint` (29), `LossOfControl` (31), `Purging` (33), and `Overexercise` (35). Notably, `ConcernDoubts` (6) also exhibited a positive association with `EatingConcern` (26). Additionally, a triad of positive edges connected `FamilyPressure` (20), `PeerPressure` (21), and `MediaPressure` (22).
- **Negative Associations (Red Edges):** Inverse relationships were identified between specific node pairs across different domains. These included associations between `PersonalStandards` (7) and Loneliness (8), and between EmotionRegulation (9) and SelfEsteem (10). Furthermore, a negative association was observed between FamilyIncome (17) and Food insecurity (19).
- **Non-Valence Associations (Grey Edges):** The network is characterized by a high density of non-valence edges, which represent dependencies where a directional sign is not mathematically defined. This occurs in associations involving categorical variables. Prominent grey edges were observed between demographic variables, specifically `Regions` (11), `Caste` (15), and `Religion` (16). Structural and biological variables also showed strong non-valence associations, such as those between `Urban` (12) and `Laxatives` (34), and `EatingDisorderParents` (5) with Food insecurity (19), Purging (33), PersonalStandards (7), and ConcernDoubts (6). Additionally, HomeTypes (18) was associated with both Food insecurity (19) and FoodSources (24), while Gender (2) was linked to Employment (14).

In summary, the weighted MGM reveals a complex architecture of dependencies, where positive associations dominate the cognitive and behavioral domains, while non-valence edges bridge socio-demographic, structural, and psychological variables.

### Network Stability

To assess the reliability of the estimated network structure and edge weights, a non-parametric bootstrapping procedure (1,000 iterations) was conducted. The resulting stabilized network, which visualizes only those edges that were consistently present across bootstrap samples, is presented in Figure 3. The stabilized network is notably sparser than the full weighted MGM, yet it reveals a highly robust set of associations primarily defined by socio-demographic, structural, and behavioral variables.

**Figure 3:**
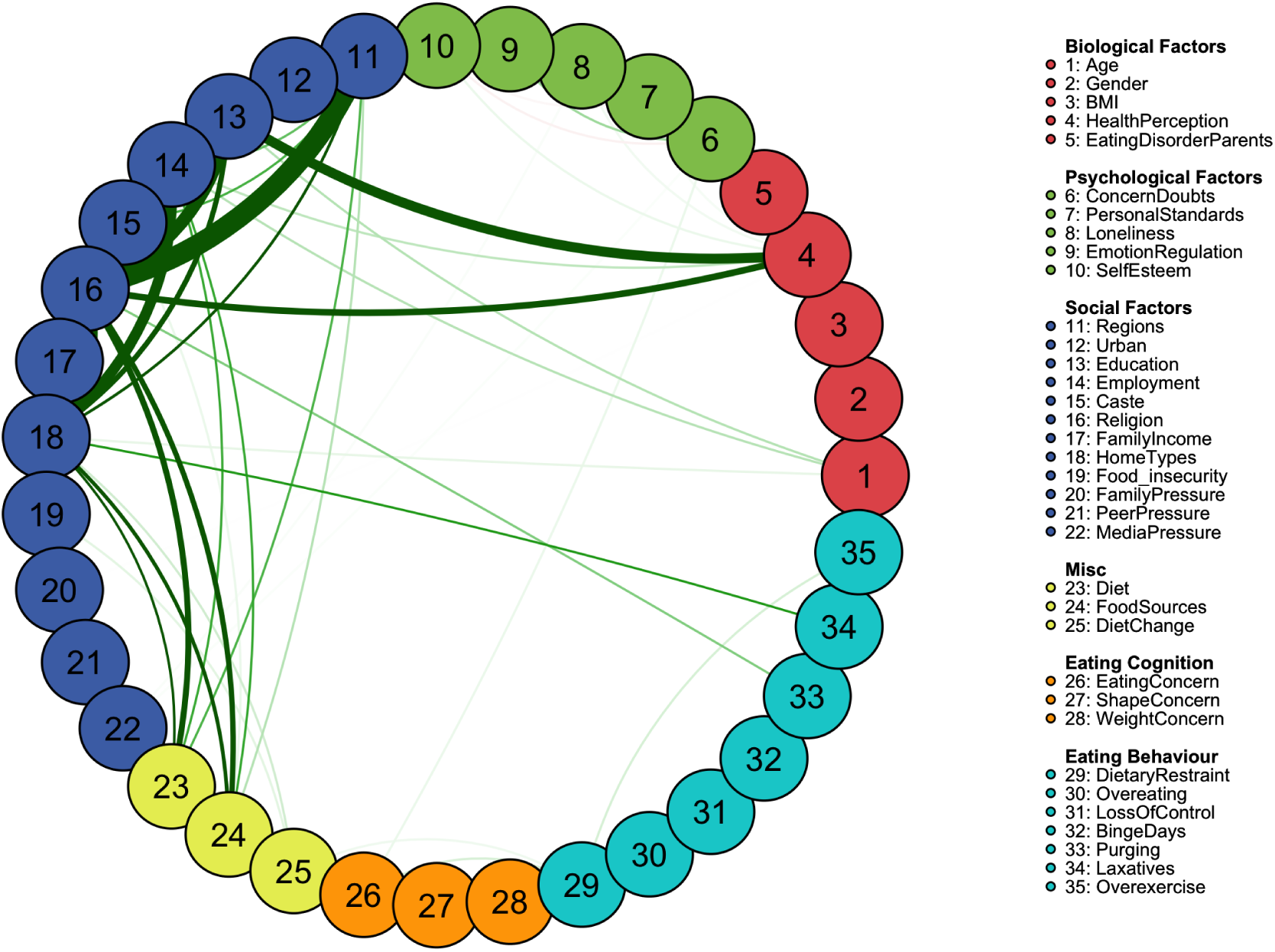
The Stabilized Network of Non-Clinical Eating Behaviors. The network visualizes edges with high stability as determined by a non-parametric bootstrapping procedure. Node colors and layout are identical to Figure 2. In this visualization, green edges represent both stable positive-valence associations and stable non-valence associations. Stable negative-valence associations (not observed in this network) would be colored red.

All stable edges identified in this procedure are represented as green in Figure 3, signifying robust conditional dependencies. The most prominent stable associations were observed between structural and environmental variables, specifically involving `Regions` (11), `Education` (13), `Employment` (14), `Religion` (16), and `HomeTypes` (18). Robust edges linked `Regions` (11) with `Religion` (16), `HomeTypes` (18), `Diet` (23), and `FoodSources` (24). Similarly, `Education` (13) showed stable associations with `Religion` (16), `HomeTypes` (18), and `HealthPerception` (4). `Employment` (14) was stably connected to `HomeTypes` (18), `Diet` (23), `FoodSources` (24), and `HealthPerception` (4). Notably, `HealthPerception` (4) also exhibited a strong stable connection with `Religion` (16).

Furthermore, the stability analysis identified reliable pathways linking these structural anchors to specific behaviors. `HomeTypes` (18) exhibited stable edges with `Diet` (23), `FoodSources` (24), and `Laxatives` (34). `Religion` (16) was stably associated with `Diet` (23), `FoodSources` (24), and `Purging` (33). Other consistently observed edges included the influence of `Age` (1) on `Education` (13) and `Employment` (14), and within the psychological domain, the edge between `PersonalStandards` (7) and `Loneliness` (8) remained stable.

In summary, while the full MGM identifies a dense web of interactions, the stability analysis highlights that the most reliable conditional dependencies in this Indian cohort are those anchoring individual behaviors, health perceptions, and psychological states to broader socio-demographic and structural contexts.

### Global Network Topology Measures & Small-worldness

The network exhibited a high Global Clustering Coefficient (Transitivity) of 0.583 and a low Average Weighted Distance (L) of 0.030. This combination of high local density and short global connectivity is a hallmark of a “small-world” architecture. The presence of this structure was confirmed by an exceptionally high Small-worldness Index (S=54.64), indicating that the Indian eating behavior landscape is significantly more optimized for rapid, system-wide integration than a random network of the same size and degree distribution.

Furthermore, the network was found to be disassortative (Assortativity = -0.143). This negative correlation suggests a “star-like” topology where highly central hubs (such as `Employment` and `HomeTypes`) tend to connect to less central, more peripheral nodes. In a public health context, this disassortative small-world structure implies a system that is robust to random fluctuations but highly vulnerable to targeted disruptions of its primary hubs.

### Community Detection Analysis (Segregation)

The Louvain algorithm identified a 6-community solution, revealing a highly organized functional landscape where variables cluster based on their mutual conditional dependencies (Figure 4).

- **Community 1 (Demographic-Labor):** This module represents the intersection of life-stage and social participation, clustering `Age` (1), `Gender` (2), `Education` (13), and `Employment` (14).
- **Community 2 (Eating Pathology Core):** This large community integrates biological markers with clinical concerns and behaviors. It contains BMI (3), the cognitive triad of EatingConcern (26), ShapeConcern (27), and WeightConcern (28), and behaviors including DietaryRestraint (29), LossOfControl (31), BingeDays (32), and Overexercise (35).
- **Community 3 (Psychological Resilience & Vulnerability):** This module captures the internal psychological landscape, clustering `HealthPerception` (4), `ConcernDoubts` (6), `PersonalStandards` (7), `Loneliness` (8), `EmotionRegulation` (9), and `SelfEsteem` (10).
- **Community 4 (Socio-Environmental Risk):** This community highlights the role of the physical and economic environment, grouping `EatingDisorderParents` (5), `Urban` (12), `FamilyIncome` (17), `HomeTypes` (18), `Food insecurity` (19), `FoodSources` (24), `DietChange` (25), and `Laxatives` (34).
- **Community 5 (Cultural-Behavioral Interface):** This distinct module links structural-cultural identity with specific dietary and compensatory practices. It includes `Regions` (11), `Caste` (15), `Religion` (16), and `Diet` (23), alongside `Overeating` (30) and `Purging` (33).
- **Community 6 (Social Pressure):** A localized module focused exclusively on external interpersonal influences: `FamilyPressure` (20), `PeerPressure` (21), and `MediaPressure` (22).

**Figure 4:**
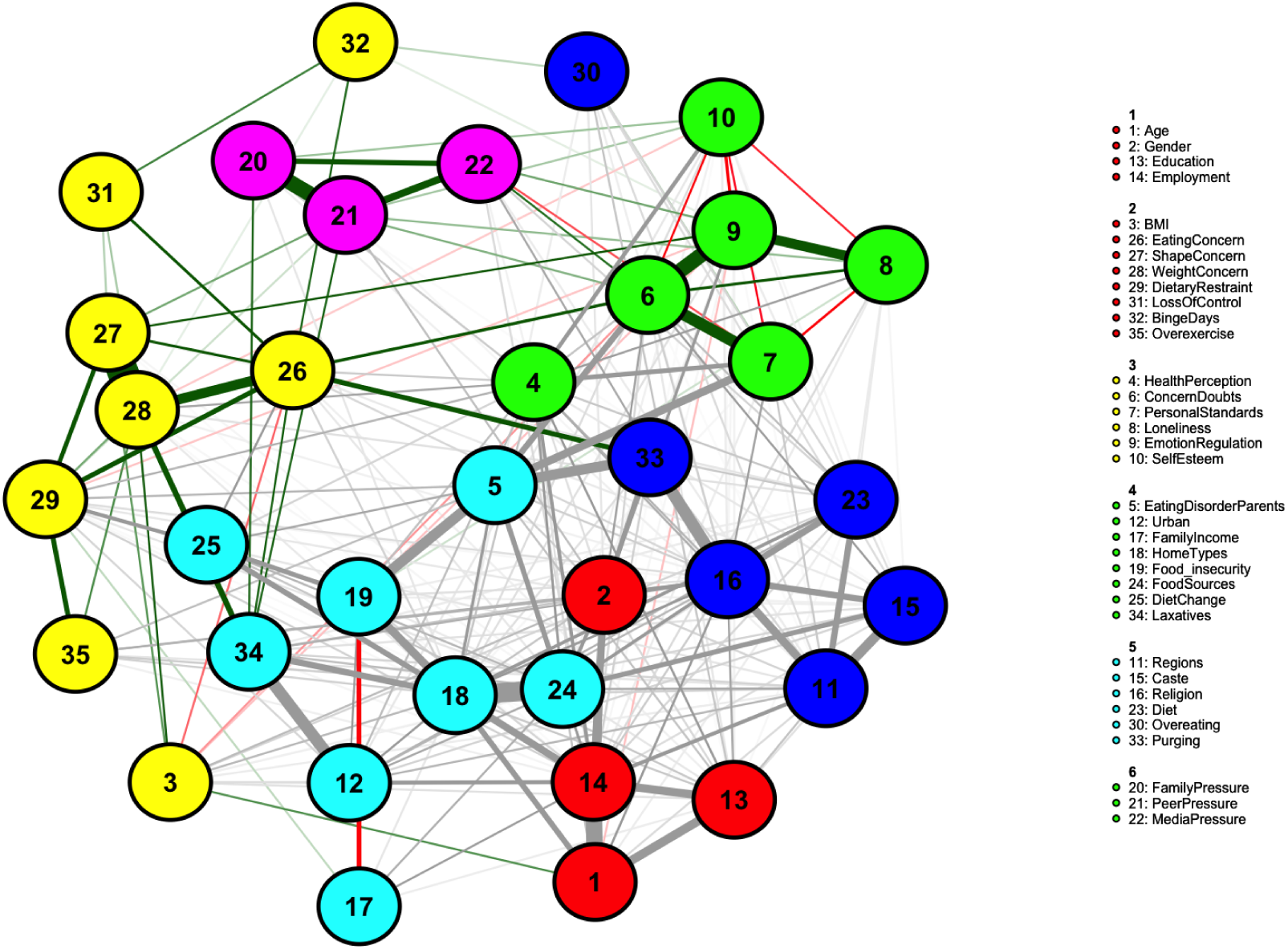
Community Structure of the Biopsychosocial Network. The network identifies a 6-community structure using the Louvain method. Node colors represent their assigned functional modules: Demographic-Labor (C1), Eating Pathology Core (C2), Psychological Resilience & Vulnerability (C3), Socio-Environmental Risk (C4), Cultural-Behavioral Interface (C5), and Social Pressure (C6).

These findings demonstrate that the Indian eating behavior landscape is not a homogenous web of correlations but is organized into specialized “functional neighborhoods.” The presence of variables like `Caste` and `Religion` within a community that includes `Purging` and `Diet` suggests that in this population, specific disordered behaviors may be uniquely anchored in cultural and regional frameworks rather than purely psychological ones.

### Graph Theoretic Centrality Measures (Integration)

Centrality measures (Figure 5) revealed a dual-layered hierarchy :

- **Expected Influence (EI):** The analysis revealed that the most influential nodes in the network are primarily structural and cultural. The highest EI was observed for `HomeTypes` and `Religion`, followed by `Employment` and `FoodSources`. A secondary tier of influence was observed for `EatingDisorderParents`, `ConcernDoubts`, `HealthPerception`, and `Gender`. This suggests that the overall “energy” or state of the network is most strongly governed by these environmental and cultural anchors.
- **Betweenness Centrality:** A distinct hierarchy emerged for this metric, led by socio-economic and psychological regulators. The highest betweenness was observed for `Employment`, Education, and SelfEsteem. These were followed by Caste, EmotionRegulation, DietaryRestraint, Region, and Religion.

**Figure 5:**
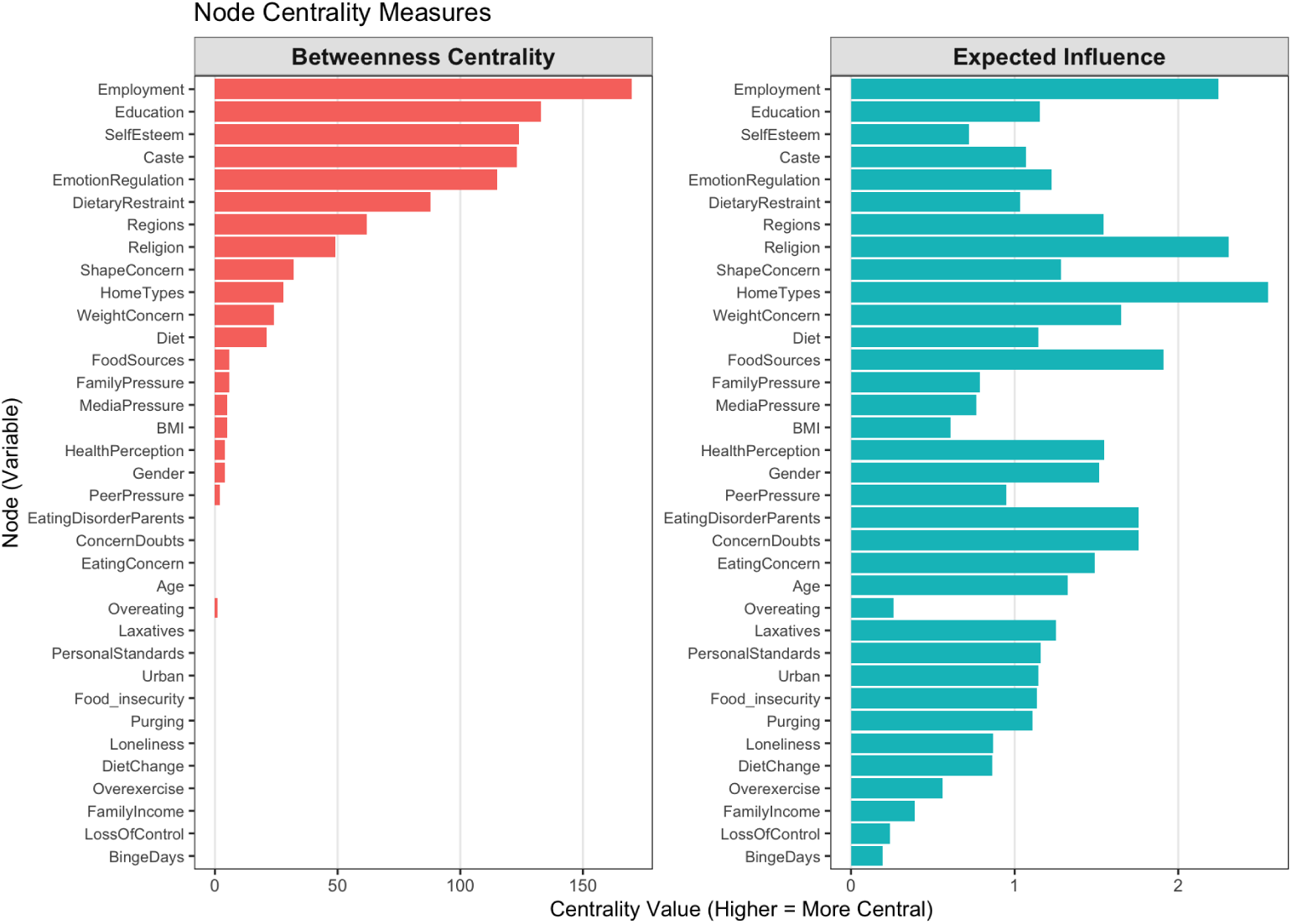
Standardized Centrality Measures for all 35 Nodes. The plot displays standardized z-scores for Expected Influence (EI) and Betweenness (bridge) centrality. Nodes are ordered by their assigned domains (color-coded at the bottom) for clarity. Key socio-economic and psychological nodes demonstrate the highest centrality across both metrics.

In summary, the centrality analysis reveals a dual-layered architecture of integration. While `HomeTypes` and `Religion` serve as the primary sources of structural influence (EI), the global connectivity of the system is maintained by a “regulatory bridge” of socio-economic (`Employment`, `Education`) and psychological (`SelfEsteem`, `EmotionRegulation`) nodes. These brokers provide the pathways through which cultural and environmental factors translate into specific behaviors, such as DietaryRestraint.

## Discussion

This study provides a systems-level map of the biopsychosocial landscape of non-clinical eating behaviors in India, revealing a network defined by a fundamental tension between modular segregation and systemic integration. In brief, (1) the network showed a highly optimized, small-world architecture; (2) cultural and living-condition variables (religion, home type) were the strongest local anchors, while socioeconomic variables (employment, education) and self-esteem served as the structural bridges integrating the whole system; and (3) shape and weight concern, though highly predictable, functioned as local rather than global integrators. We unpack each of these in turn below. Our central finding is that while the network partitions into distinct, specialized communities—ranging from a Demographic-Labor module to a Cultural-Behavioral Interface—this segregation is bridged by a small set of highly central “brokers.”

Specifically, we identify a dual-layered hierarchy of importance within the network. While structural and cultural variables—notably `HomeTypes` and `Religion`—exert the highest Expected Influence, serving as the primary anchors of the network’s local state, the global integration of the entire system is maintained by `Employment`, `Education`, and `Self-Esteem`. These nodes function as the critical “bridges” (high Betweenness) that link disparate modules. Following the “extended network” framework (De Boer et al., 2021), these results suggest that the psychological landscape of eating behavior is not an isolated internal process but is distributed across a system where environmental and structural factors play a fundamental role. The high centrality of `HomeTypes` implies that the physical and social organization of the household represents a foundational “upstream” context associated with “downstream” behavioral patterns.

This architecture profoundly reframes the discourse on non-clinical eating behaviors in the Global South. It provides quantitative evidence that these vulnerabilities are not isolated “psychological” problems but are deeply interwoven with the structural scaffolds of an individual’s life. The high betweenness of `Employment` and `Education` suggests they are not merely passive demographic markers; they function as active “highways” through which cultural and environmental pressures relate to behavioral phenotypes. Their role as stable anchors is further reinforced by their high predictability, indicating they are strongly governed by their neighboring nodes within the system. Similarly, the co-dominance of `Self-Esteem` and `Emotion Regulation` as bridges aligns with transdiagnostic models, positioning them as the core regulatory mechanisms that integrate external socio-economic contexts with internal affective states.

Our results in part support, and in part critically extend, the existing network-based literature on eating disorders and related psychopathology. Our results reveal a network characterized by high “small-worldness” (Table 3), a topological property often found in psychopathological networks which indicates a system optimized for rapid communication between nodes (Watts and Strogatz, 1998; Robinaugh et al., 2020). In practical terms, this suggests that in the Indian context, stressors can rapidly propagate from structural environments to individual psychological states.

However, our works offers a critical, culturally-specific corrective to “universalist” models derived from Western, Educated, Industrialized, Rich, and Democratic (WEIRD) populations (Henrich et al., 2010; Portingale et al., 2024). While Western literature identifies body image concerns as the primary engine of eating pathology (Fairburn et al., 2003a; Rosen, 2013; Ahrberg et al., 2011), our Indian network suggests a different hierarchy. Although Shape Concern and Weight Concern were highly predictable and influential within their local community, they did not function as the primary integrators of the global system. Instead, their influence was subordinate to the overwhelming centrality of structural factors.

This pattern directly echoes the conceptual critique raised by Monteleone and Cascino (2021), who observed that the apparent centrality of body-image cognitions in eating disorder networks is often an artifact of narrow, body-image-centric node selection. To overcome this limitation, Monteleone and Cascino (2021) called for broadening network architectures to incorporate non-eating-specific psychopathology and “external field” environmental determinants. By responding to this recommendation in a large non-clinical Global South sample—embedding eating behaviors alongside a comprehensive suite of structural, cultural, and psychological nodes—our findings provide empirical validation for this expanded framework. Specifically, we demonstrate that while shape and weight concerns retain high local centrality within eating-specific modules, global systemic integration is driven by upstream structural determinants (such as employment and education) and core self-regulatory constructs.

This implies that in a developing economy, foundational stressors related to socio-economic stability and living conditions form the primary scaffold upon which psychological and eating-related distress is built. In this context, “downstream” concerns like body image can only be fully understood when accounted for by these “upstream” social determinants of health.

### Limitations

Several limitations warrant consideration. The cross-sectional nature of the data precludes causal inference; we cannot determine, for instance, if the stable link between Emotion Regulation and Self-Esteem represents self-esteem building regulation skills or vice versa. Longitudinal studies are required to untangle these dynamic feedback loops (Borsboom, 2017). Furthermore, while the bootstrapping procedure confirmed the robust “backbone” of structural edges (e.g., the links between Religion, Diet, and Purging), many weaker edges were sample-dependent. Third, while the inclusion of caste was a vital step toward cultural specificity, the psychometric properties of Western-derived measures in Indian populations require ongoing validation. Fourth, the sample was predominantly urban (86.4%), limiting generalizability to rural populations. Finally, a potential consideration is the absence of an international or non-Indian comparison cohort. However, network analysis is fundamentally an intra-systemic method designed to estimate conditional dependencies among variables within a target sample to reveal how a complex system self-organizes (Borsboom and Cramer, 2013; Haslbeck and Waldorp, 2020). The internal validity of such a structural map depends not on group divergence, but on the statistical reliability and stability of the estimated network—a requirement we rigorously demonstrated through non-parametric bootstrapping. Furthermore, because our 35-node network explicitly incorporates culturally anchored socio-structural variables specific to India (e.g., Caste, regional variations, localized home environments), an external non-Indian sample would lack an identical node matrix, rendering direct network comparison mathematically and conceptually invalid. Evaluating whether specific bridge nodes operate differently across distinct cultural contexts remains a compelling direction for future comparative work; however, the present study establishes the unique structural architecture of eating behaviors within this population using established WEIRD-derived literature as a conceptual benchmark.

### Implications

These limitations notwithstanding, our findings open several clear avenues for future research. The most pressing is the need for longitudinal studies to map the temporal evolution of this network, especially during key life transitions such as entering or exiting the educational system or the workforce. A second, more exciting, avenue lies in intervention. Our results provide a clear empirical rationale for a two-pronged approach. Public health strategies may be most effective not by narrowly targeting ‘eating behaviors,’ but by addressing the structural integrators: `Education` (13) and `Employment` (14). This could involve programs that integrate mental health support into educational institutions or vocational training centers. Concurrently, at the individual level, our findings strongly advocate for transdiagnostic, resilience-building interventions focused on the `SelfEsteem` (10) hub. Such interventions could theoretically act as a “firewall,” buffering the individual from network-wide cascades of distress. Future research could also use our model for comparative network analysis—for example, contrasting the network structures of urban versus rural, or male versus female populations, to understand differential vulnerabilities.

In conclusion, this analysis demonstrates how a complex biopsychosocial system self-organizes in the Indian context. By moving beyond a simple list of risk factors, we show that eating behaviors are part of a structured ecosystem linked by powerful socio-economic and psychological hubs. This work challenges the primacy of body-image-centric models in the Global South and provides a data-driven blueprint for designing more effective, culturally-attuned, and systemic interventions in India.

## Data Availability

The dataset analyzed during the current study is not publicly available due to privacy and ethical restrictions regarding sensitive eating behavior and psychological data, but is available from the corresponding author upon reasonable request. The complete R scripts used for data preprocessing, Mixed Graphical Model (MGM) estimation, and network analysis are openly available in the GitHub repository at https://github.com/dipanjan-neuroscience/EatingBehaviorNetwork/

## Author Contributions

**DR** Conceptualization; Methodology; Writing – original draft; Supervision; Funding acquisition. **AR** Conceptualization; Investigation; Writing – review & editing. **EC** Conceptualization; Investigation; Writing – review & editing. **MD** Formal analysis; Writing – original draft; Validation; Visualization.

## Acknowledgments

The authors thank the 1,508 participants across India for their time and contribution to this study.

## Data Accessibility Statement

### Materials, Data, and Code Availability

All study materials (including the complete survey instrument and codebook), the de-identified dataset, and fully documented analysis scripts are publicly available in the GitHub repository at https://github.com/dipanjan-neuroscience/EatingBehaviorNetwork.

### Privacy Protection Measures

To safeguard participant privacy in compliance with our approved IRB protocol, the shared dataset has been anonymized as follows:

- All free-text responses have been removed or redacted.
- Timestamps and IP addresses have been completely removed.

### Repository Contents

The repository contains:

- De-identified dataset prepared for network estimation.
- Fully documented R scripts in numbered execution order covering: (a) data preprocessing and cleaning, (b) Mixed Graphical Model (MGM) network estimation, (c) bootstrap stability analysis, (d) community detection, (e) centrality calculations, and (f) figure generation for the main text and supplementary materials.
- A comprehensive `README.md` file providing step-by-step replication instructions.

### Restricted Data Access

The raw dataset containing unredacted free-text responses cannot be publicly released to protect participant privacy. De-identified raw data outside the public release parameters remain available from the corresponding author upon reasonable request, subject to a standard data use agreement and institutional ethics oversight.

## Footnotes

1 We retain the term WEIRD here because it is now well established in the psychological literature as shorthand for this specific sampling bias, with a substantial citation record (Henrich et al., 2010); we use it descriptively, to characterize the composition of prior samples, not as a value judgment about any population.

2 Here and throughout, we use “Global South” in its now-standard geopolitical sense—referring to low- and middle-income countries across Asia, Africa, and Latin America that share broadly comparable histories of colonization and development—rather than as a strictly geographic (southern-hemisphere) designation., where the structural determinants of health may play a more foundational role in shaping behavioral phenotypes Staples (2016); Dey and Sarkar (2025). Although our empirical data were collected exclusively in India, these shared systemic dynamics—including high mental health treatment gaps, structural socio-economic stressors, and relational health frameworks—provide strong grounds to believe our findings hold broader relevance across other Global South contexts.

3 We use “downstream” to refer to individual-level symptoms and behaviors (e.g., dietary restraint, body dissatisfaction) that are the typical focus of clinical assessment, and “upstream” to refer to the socio-structural conditions (e.g., household environment, employment, education) that shape and sustain those symptoms. These terms describe relative position within the network’s hierarchy of influence, not a claim of temporal or causal order.

4 Socio-political identity was categorized according to the Government of India’s classification system: General Category (historically more privileged groups), Other Backward Classes (OBC), and Scheduled Castes/Tribes (SC/ST) (groups historically facing structural and socio-economic marginalization). These are the official classifications used in Government of India policy and census data; we retain this terminology, rather than a paraphrase, for accuracy and comparability with national statistics, not to imply an evaluative judgment about any group.

