## Supplementary Document for "The Network Landscape of Non-Clinical Eating Behaviors in India"

### 1 Detailed Methods

#### 1.1 Measures

##### 1.1.1 Disordered Eating Behaviors and Cognitions

The **Eating Disorder Examination Questionnaire 6.0 (EDE-Q 6.0)** was used to assess both behavioral and psychological features of disordered eating (Fairburn and Beglin, 1994). Behavioral markers—including episodes of overeating, loss of control, binge eating, laxative misuse, purging, and over-exercise—were measured via frequency-based items (e.g., "Over the past 28 days, on how many days have episodes of overeating occurred?"). Psychological symptoms were captured through four subscales: Restraint, Eating Concern, Weight Concern, and Shape Concern. Items were rated on a 7-point Likert scale ranging from 0 (No days) to 6 (Every day).

##### 1.1.2 Emotion Regulation

The **Difficulties in Emotion Regulation Scale (DERS-16)** assessed participants' capacity to manage emotional distress across five dimensions: Clarity, Goals, Impulse, Strategies, and Non-acceptance (Bjureberg et al., 2016). Items (e.g., "When I am upset, I have difficulty thinking about anything else") were rated on a 5-point Likert scale from 1 (Almost Never) to 5 (Almost Always). Higher total scores indicate greater emotional dysregulation.

##### 1.1.3 Perfectionism

Aspects of perfectionism were measured using two subscales of the **Frost Multidimensional Perfectionism Scale (FMPS)** (Frost et al., 1990): *Perfectionistic Concerns* (e.g., "People will probably think less of me if I make a mistake") and *Perfectionistic Standards* (e.g., "If I do not set the highest standards for myself, I am likely to end up a second-rate person"). Items utilized a 5-point Likert scale (1 = Strongly Disagree to 5 = Strongly Agree).

##### 1.1.4 Self-Esteem

Global self-worth was assessed using the **Rosenberg Self-Esteem Scale (RSES-10)** (Rosenberg, 1965). The scale consists of 10 items (e.g., "On the whole, I am satisfied with myself" and "At times I think I am no good at all"). Respondents rated items on a 4-point scale from 1 (Strongly Agree) to 4 (Strongly Disagree).

##### 1.1.5 Social and Sociocultural Factors

- **Loneliness:** The 6-item **De Jong Gierveld Social Isolation Scale (DJGS)** measured emotional and social loneliness (De Jong Gierveld and Van Tilburg, 1999). Items (e.g., "I experience a general sense of emptiness") were scored categorically (Yes, More or Less, or No).
- **Sociocultural Pressure:** The **SATAQ-4** evaluated perceived pressure from Family, Peers, and Media to conform to appearance ideals (Schaefer et al., 2015). Items were rated on a

5-point scale (1 = Definitely Disagree to 5 = Definitely Agree).

- **Food Insecurity:** Access to food was measured using the **Household Food Insecurity Access Scale (HFIAS)**, where low scores indicate stable physical and economic access to food (Coates et al., 2007).

###### 1.1.6 Health and Behavioral Indicators

Self-perceived health was assessed via a single item ("What is your perception of your current state of health?"), with responses ranging from Excellent to Poor. Additionally, food sourcing was captured by identifying the primary method of daily food preparation (e.g., self-prepared, restaurants, or canteen).

##### 1.2 Formal Mathematical Specification of the MGM

#### Mixed Graphical Model (MGM)

Let $X = (X_1, X_2, \ldots, X_p)$ be a random vector where each variable $X_s$ follows a univariate exponential family distribution with sufficient statistic $\phi_s(\cdot)$ and base measure $C_s(\cdot)$. The pairwise mixed graphical model is defined by the joint distribution

$$P(X) = \exp\left\{ \sum_{s\in V} \theta_s \phi_s(X_s) + \sum_{(s,t)\in E} \theta_{st}\phi_s(X_s)\phi_t(X_t) + \sum_{s\in V} C_s(X_s) - \Phi(\theta) \right\},$$

where:

- $V = \{1, 2, \ldots, p\}$ is the set of nodes (variables),
- $E \subseteq V \times V$ is the set of undirected edges,
- $\theta_s$ is the node potential,
- $\theta_{st}$ is the edge potential,
- $\Phi(\theta)$ is the log-normalizing constant.

The conditional distribution of $X_s$ given all other variables $X_{\setminus s}$ is

$$P(X_s \mid X_{\setminus s}) \propto \exp\left\{ \theta_s \phi_s(X_s) + \sum_{t\in N(s)} \theta_{st}\phi_s(X_s)\phi_t(X_t) + C_s(X_s) \right\},$$

where $N(s) = \{t : (s,t) \in E\}$ is the neighborhood of node $s$.

#### Nodewise Estimation with Cross-Validation

The graph $G = (V, E)$ is recovered by estimating, for each node $s$, its neighborhood $N(s)$. This is done by solving an $\ell_1$-regularized generalized linear model where $X_s$ is the response and the predictors are all other variables and their pairwise interactions (up to a prespecified order $d$). Let $Z^{(s)}$ denote the design matrix constructed from the observations of the other variables and their interactions. For node $s$, we minimize

$$\min_{(\theta_{0s}, \boldsymbol{\beta}_s) \in \mathbb{R}^{q_s}} \left[ \frac{1}{n} \sum_{i=1}^{n} \ell\big(y_{i,s},\, \theta_{0s} + (Z_i^{(s)})^\top \boldsymbol{\beta}_s\big) + \lambda \|\boldsymbol{\beta}_s\|_1 \right],$$

where:

- $y_{i,s}$ is the $i$-th observation of $X_s$,
- $\ell(\cdot,\cdot)$ is the negative log-likelihood (deviance) corresponding to the exponential family of $X_s$,
- $q_s$ is the number of predictors (depends on $d$ and the levels of categorical variables),
- $\lambda \geq 0$ is the regularization parameter.

##### Selection of $\lambda$ by Cross-Validation

The optimal $\lambda$ is chosen via $K$-fold cross-validation:

1. Partition the $n$ observations into $K$ folds.
2. For each candidate $\lambda$ and each fold $k$:
   - Fit the model on the $K - 1$ training folds.
   - Compute the average deviance (prediction error) on the held-out fold $k$.
3. Average the deviance over the $K$ folds to obtain the cross-validated error curve $\mathrm{CV}(\lambda)$.
4. Select $\lambda^*$ that minimizes $\mathrm{CV}(\lambda)$ (or the largest $\lambda$ within one standard error of the minimum).

##### Thresholding and Edge Selection

After obtaining the estimated coefficient vector $\hat{\boldsymbol{\beta}}_s(\lambda^*)$, a hard threshold $\tau_n$ is applied:

$$\hat{\beta}_{s,t} = 0 \quad \text{if } |\hat{\beta}_{s,t}| < \tau_n,$$

where $\tau_n$ scales as $\tau_n \asymp \sqrt{d}\|\boldsymbol{\beta}_s\|_2\sqrt{\frac{\log p}{n}}$. This step enforces sparsity beyond the lasso.

The final graph is constructed by combining the nodewise estimates. Let $\hat{E}_s$ be the set of neighbors of $s$ indicated by nonzero entries in $\hat{\boldsymbol{\beta}}_s$. Then an edge $(s,t)$ is declared present if:

- **AND-rule:** $t \in \hat{E}_s$ and $s \in \hat{E}_t$,

- **OR-rule:** $t \in \hat{E}_s$ or $s \in \hat{E}_t$ (more liberal).

The Mixed Graphical Model (MGM) estimates the joint distribution of the variables by combining different exponential families. For a set of $p$ variables, the pairwise MGM joint density is given by:

$$P(X = x) \propto \exp\left(\sum_{s=1}^{p} \phi_s(x_s) + \sum_{s<t} \phi_{st}(x_s, x_t)\right)$$

where $\phi_s$ represents the node-specific univariate potentials and $\phi_{st}$ represents the pairwise interaction potentials.

In practice, the `mgm` package estimates these potentials through a series of $p$ conditional neighborhood regressions. For each node $X_r$, we estimate:

$$E[X_r | X_{\setminus r}] = \beta_{r,0} + \sum_{t \in \setminus r} \beta_{r,t} X_t$$

where $\beta_{r,t}$ represents the conditional dependency (edge weight) between node $r$ and node $t$.

##### 1.3 Node Predictability Measures

After estimating the network structure, we can assess the predictability of each node—i.e., how well it can be predicted by its neighboring nodes in the graph. Because the graph is estimated via nodewise regressions (each node regressed on all others), the fitted model directly provides a measure of the node's dependence on the rest of the network.

To quantify the extent to which each node is determined by its neighborhood, we calculated predictability. For continuous (Gaussian) nodes, we utilized the $R^2$ (coefficient of determination):

$$R_j^2 = 1 - \frac{\sum(x_{ij} - \hat{x}_{ij})^2}{\sum(x_{ij} - \bar{x}_j)^2}$$

For categorical and count nodes, we reported Classification Correctness ($CC$), which measures the accuracy of the neighborhood regression in predicting the node's state, normalized against the baseline marginal distribution. This node-wise predictability offers insight into how strongly each variable is embedded in the network.

##### 1.4 Network Topology and Centrality

To evaluate the structural importance of nodes within the biopsychosocial system, we calculated the following metrics:

1. **Expected Influence (EI):** Unlike traditional Strength centrality, EI accounts for the sign of the edges. It is defined as the sum of the absolute values of the edge weights, retaining the original sign of the association:

$$EI_i = \sum_{j \in V} \omega_{ij} \tag{1}$$

Nodes with high EI are considered the most influential "activators" or "inhibitors" of the network.

2. **Betweenness Centrality ($C_B$):** This measures the extent to which a node lies on the shortest paths between other nodes:

$$C_B(v) = \sum_{s \neq v \neq t} \frac{\sigma_{st}(v)}{\sigma_{st}} \tag{2}$$

where $\sigma_{st}$ is the total number of shortest paths from node $s$ to node $t$. This identifies "bridge nodes" that connect disparate clusters (e.g., connecting SES to Eating Behavior).

##### 1.5 Community detection: Louvain algorithm

To identify functional modules, we employed the **Louvain algorithm**, which maximizes the modularity $Q$:

$$Q = \frac{1}{2m} \sum_{ij} \left[ \omega_{ij} - \frac{k_i k_j}{2m} \right] \delta(c_i, c_j) \tag{3}$$

Where:

- $\omega_{ij}$ represents the weight of the edge between nodes $i$ and $j$.
- $k_i$ and $k_j$ are the sums of the weights of the edges attached to nodes $i$ and $j$, respectively.
- $m$ is the total weight of all edges in the network ($m = \frac{1}{2} \sum_{ij} \omega_{ij}$).
- $c_i$ and $c_j$ are the community assignments of nodes $i$ and $j$.
- $\delta(c_i, c_j)$ is the Kronecker delta function, which equals 1 if $c_i = c_j$ and 0 otherwise.

The Louvain method operates through an iterative two-phase process:

1. **Modularity Optimization:** Initially, each node is assigned to its own community. Nodes are then iteratively reassigned to neighboring communities that yield the maximum increase in $Q$.
2. **Community Aggregation:** Nodes belonging to the same community are consolidated into a single "super-node," creating a new, coarse-grained network.

These phases repeat until no further increase in modularity is possible, revealing the optimal hierarchical community structure.

##### 1.6 Data Pre-processing and Stability

**Data Pre-processing and Management** Data preparation and statistical analyses were conducted in `R` (v4.3.0). Prior to model estimation, the following steps were implemented:

- **Variable Transformation:** Socio-economic and demographic variables were mapped to numeric factors. `FamilyIncome` underwent a $log(1 + x)$ transformation to account for heavy right-skewness.
- **Outlier Handling:** To prevent extreme values from inflating partial correlations, we applied manual Winsorization to behavioral variables (e.g., `LossOfControl`, `BingeDays`), capping values at the 99th percentile ($x_{capped} = \min(x, P_{99})$).
- **Cleaning:** BMI values outside the physiological range ($< 10$ or $> 100$) were treated as missing. We utilized listwise deletion for missing values, as the MGM algorithm requires a complete case matrix for cross-validation.

**Stability Analysis:** We performed 1,000 non-parametric bootstrap iterations. The Centrality Stability (CS) coefficient was calculated to ensure the reliability of node rankings. A CS-coefficient $> 0.50$ indicates that node rankings are stable even when a significant portion of the sample is dropped (Epskamp et al., 2018).
